# Health and economic impact of caregiving on informal caregivers of people with chronic diseases in Sub-Saharan Africa: A systematic review

**DOI:** 10.1101/2024.05.24.24307869

**Authors:** Ephraim Kisangala, Etheldreda Leinyuy Mbivnjo, Edward JD Webb, Barbara Barrett, Godfrey Zari Rukundo, Eve Namisango, Margaret Heslin

## Abstract

**Background:** With a disproportionate burden of chronic diseases and severe shortage of health workers in sub-Saharan Africa (SSA), the region implicitly relies on informal caregivers (ICGs) to support the patients both within and outside the health facilities. The aim of this review is to explore the health and economic impact of caregiving on ICGs of patients with chronic diseases in SSA.

**Methods:** Medline (Ovid), CINAHL (EBSCOhost), PsycINFO (Ovid), Embase (Ovid), Global Health and Web of Science databases were systematically searched to identify original articles that considered the economic and/or health impacts of caregiving in SSA. The results from the included studies were synthesised narratively.

**Results:** After screening 4,372 articles, 47 were included for synthesis. The articles were from all sub-regions of SSA with more than half (25/47) of the articles focussing on caregivers for patients with cancer. Although the primary motivation for becoming caregivers was love and responsibility, the caring responsibilities described in twenty articles, had profound effects on the caregiver’s lives. Healthwise, ICGs experienced changes in their physical and mental health like developing musculoskeletal problems and depression. Economically, caregiving was expensive, and financially draining. The opportunity cost of caregiving included loss of jobs, loss of income, foregoing planned important activities and missed education opportunities.

**Conclusion:** ICGs reported a range of mainly negative health and economic effects of the work they do. Health care systems should consider how to better support ICGs in terms of their own physical and mental wellbeing and governments should consider how to better financially support ICGs.

## Introduction

Global increases in healthcare expenditure have not been proportional across the world (1). In sub-Saharan Africa (SSA), government spending on health is very low whilst out-of-pocket expenditure is significantly high. These countries do not have well-developed health insurance system and the out-of-pocket health expenditure is sometimes higher than 70% (2,3). Universal health insurance is still in its infancy in SSA, with only four countries having coverage levels above 20% (3,4). Therefore, the family (informal caregiver) of a person with a chronic disease in this region is often faced with a double burden of because of the long-term nature of the disease and the limited availability of diagnostic and disease management services (5). Chronic diseases which are projected to be the leading cause of morbidity and mortality in Africa by 2030 are slow-progressing diseases that last long in the body and cannot be transmitted from one person to another (6,7).

The term informal caregiver (ICG) is used to refer to any layperson who provides regular emotional, physical, financial, and medical support to the sick or to people with disabilities, often without expectation of an immediate reward (8,9). Throughout this review, ‘ICG’ and ‘caregiver’ are used interchangeably to mean the same thing unless otherwise specified. Many times, ICGs have a familial relationship with the care recipient, often assuming the caregiving role as parents, spouses, children, siblings, or other relatives. These caregivers are predominantly female and frequently take on the caregiving role with minimal or no preparation (10). It is understood that the ICGs in SSA perform numerous tasks, some of which are typically responsibilities of healthcare professionals in other parts of the world (10,11). They perform these activities whenever required, whether day or night, at home or in the hospital and under any situation. For example, ICGs sometimes sleep on hospital floors during the night to provide companionship and maintain close proximity to the care recipient in order to address any immediate concerns required by either the patient or the medical personnel at the hospital (12). Accordingly, they are faced with unique challenges, such as overwhelming responsibilities, high out-of-pocket health expenditure, stigma, and limited access to healthcare services (10,11).

Over the years, reviews have been conducted to assess the impact of caregiving in SSA. The focus of these reviews has been on specific conditions such as HIV (13), cancers (10), old age, end-of-life care (14), mental health disorders (15) and stroke. Other researchers limited their reviews to studies within specific countries (16) or specific settings like hospitals (17).

In this review, we aimed to systematically summarise evidence on the health and economic impact of caregiving on ICGs of patients with chronic diseases in the whole of SSA. This was achieved through the following three objectives: 1) To describe the activities ICGs of people with chronic diseases in SSA perform 2) To discuss the reasons why ICGs of people with chronic diseases in SSA took on the caregiving roles 3) To explore the health and economic impact of caregiving on ICGs of people with chronic diseases in SSA. From our understanding, this is the first review to explore how caregiving affects the health and economic aspects of caregivers of people with major chronic diseases in the whole of SSA.

## Methods

This systematic review was conducted, adhering to the guidelines laid out in the Preferred Reporting Items for Systematic Reviews and Meta-Analyses (PRISMA) statement (18). The protocol for this review was registered with PROSPERO under the registration number of CRD42022358531. The databases were searched between September and October 2022.

### Eligibility criteria

The criteria for selecting studies were as follows:

#### Study design

All original studies, irrespective of the design were included in this review. These included randomised controlled trials, case-control, cohort, cross-sectional and qualitative studies. However, commentaries and reviews were excluded.

#### Population

Studies with ICGs providing care to people with chronic diseases were included. We included the most common chronic diseases which are known to cause significant morbidity and over 80% of premature deaths - these were cardiovascular diseases, chronic obstructive pulmonary diseases, cancers and Type 2 diabetes (19). We excluded mental health conditions and those with a co-morbidity that is not a chronic disease of interest. We did not include studies that had less than half (50%) of ICGs in active caregiving roles. Caregivers may not actively provide care when the care recipient has significantly improved, died and when another person has taken over the caring role. We also excluded studies where results for the population of interest were pooled with others (such as professional caregivers, paid caregivers, and volunteers) and if the proportion of the population of interest was under 50%. Such studies would dilute the relevance of the findings and make them less applicable to ICGs, who are this review’s population of interest. Where results were presented separately for the population of interest, we included the studies regardless of what proportion of the total sample they made up.

#### Outcomes

The studies that were selected for inclusion in this review had findings with information about how caregiving affected the health or economic aspects of the ICG’s life. Such information had to be provided in the findings.

#### Setting

Studies were selected if they were conducted within SSA as defined by World Bank (https://openknowledge.worldbank.org/pages/focus-sub-saharan-africa). The search was not restricted to any language since articles published in languages other than English were translated.

### Information sources

EK searched for relevant original articles from the following databases between September and October 2022: Medline (Ovid), CINAHL (EBSCOhost), PsycINFO (Ovid), Embase (Ovid), Global Health and Web of Science. The searches were conducted from the inception of each database up to the date of each search. Full details of the searches including the databases, search strategy and the search outcomes can be found in **S1 File**.

In addition, the reference lists of selected articles and relevant systematic review reports were searched. Lastly, primary authors of editorial letters, abstracts from conference proceedings, abstracts without full-text articles and commentaries identified through the searches were contacted by email for full-text original articles.

### Search strategy

The search strategy development process involved identifying candidate search terms by reading the titles, abstracts, keywords, and search strategies in the three known related articles (17,20,21). A draft search string was formed by appropriately combining the identified search terms and MeSH terms using Boolean operators. The search terms: Informal caregiver, sub-Saharan Africa, and Chronic Diseases, were combined using the Boolean operator “AND,” while the thesauruses and MeSH words for each search term were combined using “OR.” This was improved after a review and discussion with two senior researchers (BB and MH) and a librarian from King’s College London, all of whom have experience in systematic reviews. This was purposely done to reduce the errors and increase the robustness of the search strategy. The final search string as used in each database can be found in **S1 File.**

### Selection process

The articles that were retrieved from all the searched databases were exported to Endnote (version 20) for removal of the duplicate articles. This was done by the first reviewer (EK). The remaining articles were then transferred to Rayyan software (https://www.rayyan.ai/) where title and abstract screening was done. The articles that did not meet the agreed criteria were excluded while those that potentially met the eligibility criteria were included for full-text screening. Full-text screening was carried out for the articles whose full-text versions were retrieved. The articles whose full-texts were unavailable even after contacting the primary authors were excluded. At each stage, the screening was independently conducted by two reviewers. While EK screened all the articles in both stages, the second reviewer (ELM) screened a random 20% of the articles at the title and abstract screening stage, and a random 25% during the full-text screening. There was 96% agreement between the two reviewers and all disagreements were resolved by a discussion with input from other authors. In this review, all non-English papers were translated into English using Google Translate, a choice made due to its known reliability and accuracy in English with 91% agreement between native language reviewers and reviews who use google translate (range, 85% to 97%) (22).

### Quality of the included studies

The quality of the articles was independently assessed by two reviewers (EK and ELM) using the Mixed Methods Appraisal Tool, a critical appraisal tool that has been tested and found to be reliable and efficient (23,24). It is used to assess the quality of studies of different designs including qualitative and quantitative studies. For each article (depending on the study design), there are five (5) questions that require a response of Yes, No or Can’t Tell. In this review, the Yes was given a score of 1 and No or Can’t Tell was given a score of 0. Any disagreement was resolved by consensus.

### Data extraction and analysis

Relevant data was extracted from all articles by EK and entered into a data extraction form, developed with the study objectives in mind. The data that was extracted into the form included; the name of the first author of the study, the year of publication, the country where the study was conducted, study design, sample size, participants’ age and sex, their marital and employment status, the relationship of ICG to the patient, activities done by the ICG, setting (home or hospital setting), type of chronic disease, scale used to measure impact, the health and economic impact of caregiving on ICG and the ICGs’ motivation to provide care. Thereafter, data were synthesized narratively.

## Results

### Study selection

As shown in the PRISMA diagram in Fig 1, 4372 records were retrieved through a systematic search of seven databases (4368) and a targeted search of the reference list of the already identified relevant articles (4). After removing 2005 duplicate articles, 2194 articles were excluded during the title and abstract screening and another 27 articles were also excluded because they were conference abstracts whose full-text articles could not be retrieved. During the full-text screening, 99 articles were excluded for various reasons and only 45 studies (with 47 articles) were included in the review.

**Fig 1.**
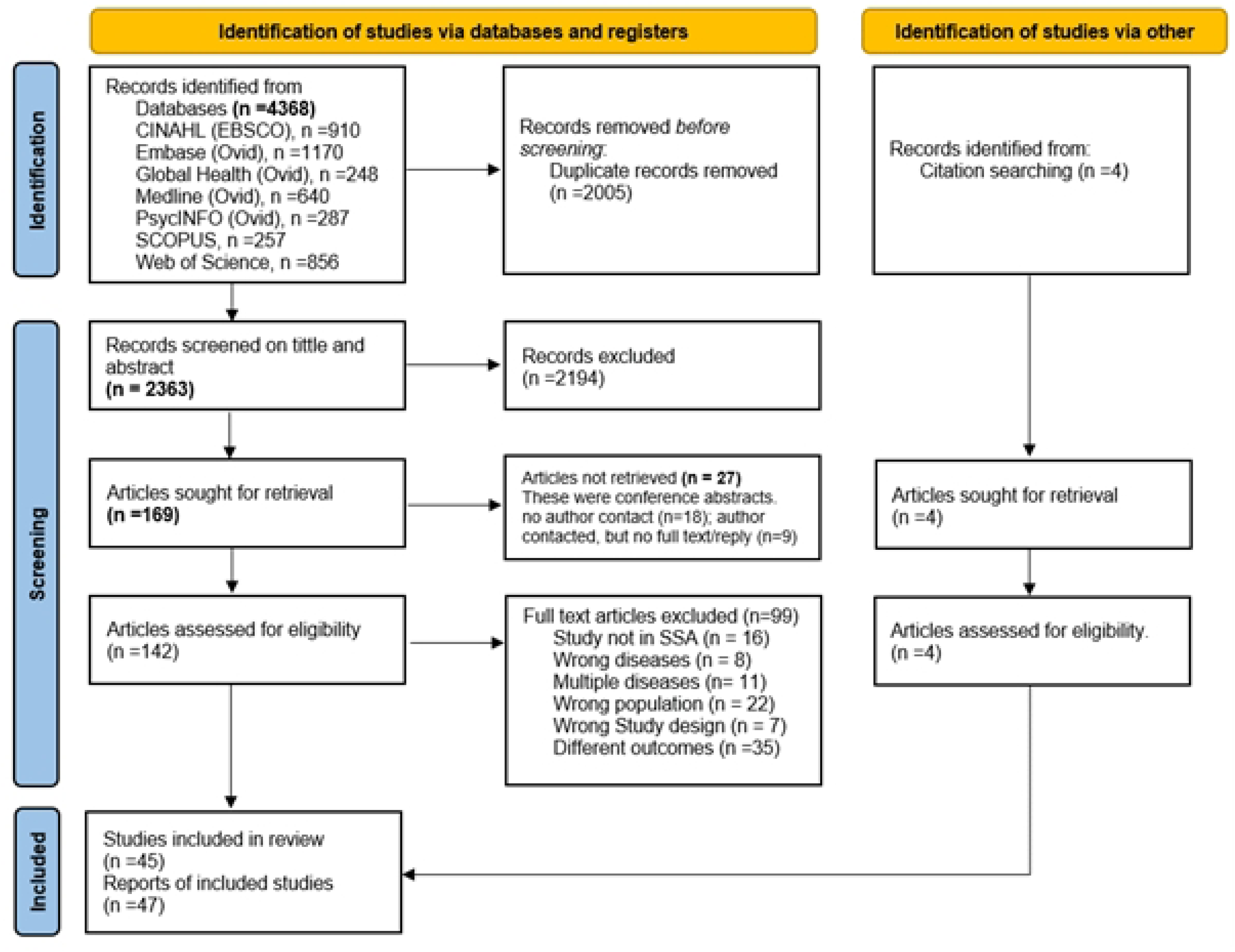
PRISMA flow diagram of the screening and selection process in this review.

### Study characteristics

All articles were written in English except one that was in French (25). The authors used qualitative and quantitative methods in 22 and 24 of the 47 articles respectively, with one article using mixed methods. Of the quantitative studies, three (26–28) were longitudinal studies and the rest had a cross-sectional study design. The articles were from fourteen countries representing all the sub-regions of SSA. The highest number of articles (11/47) were from Nigeria (29–39) and South Africa (11,26,27,40–47), while each of the following countries had one (1/47) article; Togo (48), Zimbabwe (49), Kenya (50), Sudan (51), Democratic Republic of Congo (52) and Cameroon (25). Among the articles from South Africa, two used data from the same study (42,43). Also, data from a single study in Ghana was used in two of the five articles (53,54). See Table **1**

**Table 1.** Summary of the characteristics of the reviewed articles.

| Author, year | Country | Setting | Participants | Sample size | Disease | Relationship to Patient | Duration of caring | Time spent in care |
| --- | --- | --- | --- | --- | --- | --- | --- | --- |
| Yusuf (2011)<br>(29) | Nigeria | Outpatients | Family caregivers<br>Mean age: 57 years<br>Age range: Not reported<br>Sex: Female, 43 (42%)<br>**Employment:** Unemployed 25 (24.3%)<br>Employed 78 (75.7%) | 103 | Cancers (Not specified) | Spouse: 3 (2.9%)<br>Son/daughter: 74 (71.9%)<br>Sibling: 26 (25.2%) | Not reported | Not reported |
| Wassie (2022)<br>(56) | Ethiopia | Admitted in hospital | ICGs of adults<br>Mean age: 35 years<br>Age range: Not reported<br>Sex: Female, 176 (42.72%)<br>**Employment:** Employed 282 (68.44%)/Unemployed 130 (31.56%) | 412 | Breast cancer 91 (22.09%);<br>Cervical cancer 54(13.11%);<br>Gastric cancer 30(7.28%);<br>Others 237 (57.52%) | Spouse 140 (33.98%);<br>Child 150 (36.41%);<br>Siblings 85 (20.63%);<br>Parents 21 (5.1%);<br>Other relatives 16 (3.88%); | more than 2 weeks | Not reported |
| Walubita (2018)<br>(12) | Zambia | Admitted in hospital | **ICGs of children with cancers**<br>Mean age: Not reported<br>Age range: 20 to 58 years<br>Sex: Female, 14 (93.3%)<br>**Employment:** Not reported | 15 | Cancer of the kidney (4),<br>Leukaemia (3), Retinol blastoma (3), Lymphoma (2), Cancer of the liver (1), Cancer of the lungs (1) | Parent 12;<br>Grandparent 2;<br>Great grandparent 1; | Not reported | Not reported |
| Vincent-Onabajo (2018)<br>(30) | Nigeria | Not institutionalised | ICGs of people with stroke<br>Mean age: 33.2 years<br>Age range: 19 to 60 years<br>Sex: Female, 55 (61.1%)<br>**Employment:** Unemployed 60 (66.7%)/Employed 30 (33.3%) | 90 | Stroke | Offspring 53 (58.9%);<br>Spouse 22 (24.4%);<br>Sibling 15 (16.7%). | Not reported | Not reported |
| Thomas (2008)<br>(40) | South Africa | Not institutionalised | Primary ICGs<br>Mean age: 47.2 years<br>Age range: 16 to 67 years<br>Sex: Female, Female 4 (66.7)<br>Employment: Not reported | 6 | Stroke | Husbands 2 (33.3);<br>Wife 2 (33.3%);<br>Daughters 2 (33.3%) | Not reported | Always with the care recipient |
| Tchokote (2020)<br>(25) | Cameroon | Not institutionalised | Primary or secondary ICGs<br>Mean age: 32.75 years<br>Age range: 20 to 50 years<br>Sex: Female, Female 2 (50%)<br>Employment: Employed 1 (25%); Unemployed 3 (75%) | 4 | Stroke | Grandchildren 2 (50%);<br>Wife 1 (25%); Daughter 1 (25%) | Not reported | Not reported |
| Serfontein (2019)<br>(26) | South Africa | Admitted in a Rehabilitation centre | Primary ICGs<br>Mean age: Not reported<br>Age range: 23 to 76 years<br>Sex: Female, Female 50 (79.4%)<br>Employment: Employed 30 (47.6%);<br>Unemployed 33 (52.4%) | 63 | Cerebrovascular accident (CVA) | Spouses 32 (50.8%);<br>Children 18 (28.6%);<br>Other relatives (13 20.6%) | ≥ 2 months | 13.2% (5) spend ≥4 hours/day<br>55.6% (35) spend 2-4 hours/day &<br>23.8% (15) spent 5-6 hours/day |
| Scheffler (2019)<br>(27) | South Africa | Not institutionalised | **ICGs [73/80 (91.2%)-ICGs; & 7/80 (8.8%) paid carers]**<br>Mean age: 47.4 years, | 89 | Stroke | Spouse/partner 25 (28.4%)<br>Son/daughter 33 (37.5%) | Not reported | Not reported |
|  |  |  | Age range: 12 to 79 years<br>Sex: Female, Female 73 (82.0%)<br>Employment: Not reported |  |  | Other family 19 (21.6%)<br>Friend 6 (6.8%)<br>Other 6 (5.7%) |  |  |
| Owoo (2022) (54) | Ghana | Not clear | Family caregivers<br>Mean age: 48.67 years<br>Age range: 27 to 67 years<br>Sex: Female, Female 11 (91.67%)<br>Employment: Employed: 9 (75%); Unemployed: 3 (25%) | 12 | Prostate cancer | Father 6 (50%)<br>Husband 6 (50%) | 1 year 8 (66.67%)<br>2 years 3 (25%)<br>6 months 1 (8.33%) | Not reported |
| Owoo (2022) (53) | Ghana | Not reported | Family caregivers<br>Mean age: 48.67 years<br>Age range: 27 to 67 years<br>Sex: Female, Female 11 (91.67%)<br>Employment: Employed: 9 (75%); Unemployed: 3 (25%) | 12 | Prostate cancer | Father 6 (50%)<br>Husband 6 (50%) | 1 year 8 (66.67%)<br>2 years 3 (25%)<br>6 months 1 (8.33%) | Not reported |
| Onyeneho (2021) (31) | Nigeria | Not reported | Primary caregiver<br>Mean age: 37.68 YEARS<br>Age range: 10 to >51 years<br>Sex: Female, Female 11 (91.67%)<br>Employment: Employed: 9 (75%); Unemployed: 3 (25%) | 182 | Cancer (Not specified) | Brother 12 (6.6%)<br>Sister 20 (11.0%)<br>Parents 119 (65.4%)<br>Husband 17 (9.3%)<br>Wife 14 (7.7%) | <1 month 52 (28.6%)<br>1 month to 6 months 63 (34.6%)<br>>6 months to 1 year 21 (11.5%)<br>>1 year 46 (25.3%)<br>Mean±SD 2.34±1.14 | Not reported |
| Okeke (2020) (32) | Nigeria | within outpatient department of hospital | Informal caregivers<br>Mean age: 41.1 years<br>Age range: 18 to 75 years<br>Sex: Female, Female 179 (57.6%)<br>Employment: Not reported | 311 | Stroke | Parents 100 (32.1%)<br>Children 18 (5.8%)<br>Spouse 106 (34.1%)<br>Non relatives 18 (5.8%)<br>Other relatives 69 (22.2%) | 1 to 3 months 112 (36%)<br>4 to 6 months 26 (8.3%)<br>7 to 12 months 22 (7.1%)<br>13 to 24 months 32 (10.3%)<br>More than 24 months 119 (38.3%) |  |
| Ohaeri (1999) (33) | Nigeria | Outpatient | Informal caregivers<br>Mean age: 35.6<br>Age range: 18 to 75 years<br>Sex: Not reported<br>Employment: Employed 52 (70.4%), Unemployed 21 (29.6%); | 73 | Cervix cancer<br>Breast cancer | Husband 24 (32.9%)<br>Child 36 (49.3%)<br>Others: 13 (17.8%) | Not reported | Daily contact with patient 55 (75.3%) |
| Ogunmodede (2019) (34) | Nigeria | Not institutionalised | Primary caregivers [1(1%) paid carers, 99 (99%) ICGs]<br>Mean age: 46.5 years<br>Age range: ≤20 to >60 years<br>Sex: Female 56 (56.0%) | 100 | Type 2 Diabetes Mellitus | Parents 10 (10.0%)<br>Child 40 (40.0%)<br>Spouse 34 (34.0%)<br>Siblings 11 (11.0%)<br>Unrelated 1 (1.0%)<br>Others 4 (4.0%) | Not reported | Daily contact 83 (83.0%)<br>Weekly contact 10 (10.0%)<br>2-3 times a month 5 (5.0%)<br>Several times a year 2 (2.0%)<br>**Number of contact hours**<br><= 5hours: 4 (4.0%) |
|  |  |  | Employment: Employed 66 (66.0%),<br>Unemployed 34 (34.0%) |  |  |  |  | 6-10 hours: 18 (18.0%)<br>11-15 ours 22 (22.0%)<br>>16 hours 31 (31.0%) |
| Muriuki (2023)<br>(50) | Kenya | Outpatient<br>clinic | Family caregivers<br>Mean age: Not reported<br>Age range: 18 to >61 years<br>Sex: Female 147 (57.6%)<br>Employment: Employed 109 (42.8%),<br>Unemployed 146 (57.3%) | 255 | Cancer (not specified) | Not reported | at least 2 weeks | Not reported |
| Muliira (2019)<br>(57) | Uganda | Mixed (both out<br>and inpatients) | Family caregivers<br>Mean age: 36 years<br>Age range: 18 to >51 years<br>Sex: Female 208 (73.2%)<br>Employment: Employed 161 (56.7%),<br>Unemployed 123 (43.3%) | 284 | Breast cancer (18.3%), cervix<br>cancer (12.3%), and leukaemia<br>(12.3%) and others (cancers of<br>oesophagus, prostate, ovary,<br>pancreas, stomach,<br>Lymphomas, and multiple<br>myeloma). | Spouse 57 (20.1%)<br>Child 110 (38.7%)<br>Others 117 (41.2%) | 7–11 months<br>135 (47.5%)<br>12–24 months<br>91 (32.1%)<br>≥25 months 58<br>(20.4%) | ≤48 hours/week 57 (20.1%)<br>49–120 hours/week 92 (32.4%)<br>≥121 hours/week 135 (47.5%) |
| Mthembu<br>(2016) (41) | South<br>Africa | Not reported | Informal caregivers<br>Mean age: 58.5 years<br>Age range: 24 to 79 years<br>Sex: Female 5 (83.33%)<br>Employment: Not reported | 6 | Lung Disease<br>Hypertension<br>Stroke<br>Diabetes mellitus | Spouse 2 (33.33%)<br>Children 3 (50%)<br>Other relatives<br>(Daughter in Law) 1<br>(16.67%) | Not reported | Not reported |
| Mlaba (2021)<br>(43) | South<br>Africa | Mixed | Informal caregivers (*patients of 3 (15%) of IGCs*<br>*had died*)<br>Mean age: 55.05 years<br>Age range: 21 to 84 years<br>Sex: Female 14 (70.0%)<br>Employment: Not reported | 20 | Various cancers (Not specified) | Spouse 7 (35.0%)<br>Offspring/children 5<br>(25.0%)<br>Parent 1 (5.0%)<br>Other relatives 7<br>(35.0%) | Not reported | Not reported |
| Mlaba (2020)<br>(42) | South<br>Africa | Mixed | Informal caregivers (*patients of 3 (15%) of IGCs*<br>*had died*)<br>Mean age: 55.05 years<br>Age range: 21 to 84 years<br>Sex: Female 14 (70.0%)<br>Employment: Not reported | 20 | Various cancers (Not specified) | Spouse 7 (35.0%)<br>Offspring/children 5<br>(25.0%)<br>Parent 1 (5.0%)<br>Other relatives 7<br>(35.0%) | Not reported | Not reported |
| Mensah (2021)<br>(66) | Ghana | Not reported | Husbands of women with advanced breast<br>cancer<br>Mean age: Not reported<br>Age range: 34 to 55 years<br>Sex: Female 0 (0%) Male 15 (100%)<br>Employment: Employed 15 (100%) | 15 | Breast cancer | Husband 15 (100%) | 4 to 11 months | Not reported |
| Mekonnen<br>(2020) (61) | Ethiopia | Not reported | Parents of children with cancer<br>Quantitative<br>Mean age: Not reported<br>Age range: 18 to >49 years<br>Sex: Female 145 (52.7%)<br>Employment: Not reported<br>Qualitative<br>Mean age: Not reported | 295<br>(Quant<br>itative<br>= 275;<br>Qualit<br>ative =<br>20) | Cancer (Not specified) | Quantitative Parents<br>275 (100%);<br>and<br>Qualitative<br>Parents 20 (100%) | Qualitative<br>(n=275)<br>1-3 weeks 14<br>(5.1%)<br>4-6 weeks 148<br>(53.8%)<br>7-9 weeks 64<br>(23.3%)<br>≥10 weeks 49<br>(17.8%) | Not reported |
|  |  |  | Age range: 36-45 years<br>Sex: Female 13 (65%)<br>Employment: Not reported |  |  |  |  |  |
| Masuku, (2018)<br>(44) | South Africa | Home | Primary caregivers<br>Mean age: 38 years.<br>Age range: 21 to 65 years<br>Sex: Female 14 (100%)<br>Employment: Employed 6 (42.9%), Unemployed 8 (57.1%) | 14 | stroke | Children 5 (35.7%)<br>Other relatives 1 (7.1%)<br>Wife 6 (42.9%)<br>Sister 2 (14.3%) | RANGE (3 months to more than 3 years)<br><= 6 months 6 (42.9%)<br>7 months to <= 12 months 3 (21.4%)<br>>12 months to <= 24 years 3 (21.4%)<br>24 months to 48 months 2 (14.3%) | Not reported |
| Masika (2020)<br>(62) | Tanzania | Admitted | Guardians of children<br>Mean age: 38 years<br>Age range: Not reported<br>Sex: Female 15 (68.2%)<br>Employment: Not reported | 22 | leukemia (5), eye cancer (5), cancer of the glands (5), kidney cancer (2), stomach cancer (1), thyroid cancer (1), and liver cancer (1) | Guardian 22 (100%) | Not reported | Not reported |
| Marima (2019)<br>(49) | Zimbabwe | Outpatients | ICGs<br>Mean age: 41.5 years<br>Age range: Not reported<br>Sex: Female 50 (70.4%)<br>Employment: Not reported | 71 | Stroke | Not reported | Median (IQR) 4 (2-13) months; Most had provided care for at least 4 months | Not reported |
| Maree (2018)<br>(11) | South Africa | Not reported | Primary ICGs<br>Mean age: 41.7 years<br>Age range: 20 to 65 years<br>Sex: Female 10 (50%)<br>Employment: Not reported | 20 | various cancers (Not specified) | Children: 10 (50%)<br>Spouse: 4 (20%)<br>Friends/non-related: 3 (15%)<br>Other relatives: 2 (10%)<br>Sibling: 1 (5%) | Between 3 weeks and 10 years, with an average of 24.8 months (SD ± 26.9). | Not reported |
| Kitoko (2022)<br>(52) | Democratic Republic of Congo | Outpatients (follow-up visit to rehabilitation centre) | ICGs<br>Mean age: 42.3 years<br>Age range: Not reported<br>Sex: Female 58 (68.2%)<br>Employment: Employed: 54 (63.5%); Unemployed: 31 (36.5%) | 85 | Stroke | Not reported | At least 3 months | Not reported |
| Khondowe (2007) (68) | Zambia | Outpatients | Caregivers of stroke outpatients | 10 | Stroke | Not reported | Not reported | Not reported |
| Ketlogetswe (2022) (45) | South Africa | Hospice admitted patients | Primary ICGs<br>Mean age: 52 years<br>Age range: 23 to 84 years<br>Sex: Female 19 (86.36%)<br>Employment: Employed: 10 (45.45%); Unemployed: 12 (54.54%) | 22 | cancers of head, neck, prostate, thyroid, breast, lung, vulva, & blood | mostly mothers of the sick person, but also included partners, sons, daughters and a neighbour (No statistic given) | 13 months on average | Not reported |
| Katende (2017) (60) | Uganda | inpatients and outpatients | Family caregivers<br>Mean age: 33 years | 119 | Cancer (Not specified) | Parent: 32 (26.9%)<br>Other 1st degree | Not reported | Not reported |
|  |  |  | Age range: 18 to 60 years<br>Sex: Female 80 (67.2%)<br>Employment: Employed: 73 (61.3%)<br>Unemployed: 46 (38.7%) |  |  | relatives: 49 (41.2%)<br>Extended family: 32 (26.9%)<br>Other: 6 ( 5.0%) |  |  |
| Jones (2012) (28) | Tanzania | At their home | Primary ICGs of physically dependant stroke survivor<br>Mean age: 47 years<br>Age range: 16 to 73 years<br>Sex: Female 23 (85.19%)<br>Employment: Employed: 17 (63.00%),<br>Unemployed: 10 (37.00%) | 27 | Stroke | Children: 14 (51.85%)<br>Spouse: 11 (40.74%)<br>Grand children: 2 (7.41%) | At least 6 months | Not reported |
| Gertrude (2019) (21) | Uganda | admitted in hospital | ICGs<br>Mean age: Not reported<br>Age range: 18 to ≥55 years<br>Sex: Female 14 (56%)<br>Employment: Employed: 21 (84%),<br>Unemployed: 4 (16%) | 25 | STROKE:<br>Ischaemic stroke: 15 (60%)<br>Haemorrhagic stroke: 8 (32%)<br>Stroke (type Missing): 2 (8%) | Spouse: 9 (36%)<br>Child/Child in law: 9 (36%)<br>Other: 7 (28%) | 4 months to 1 year: 10 (40%)<br>≥1 year: 15 (60%)<br>(Median duration of care: 16 months) | Not reported |
| Gbiri (2015) (35) | Nigeria | Physiotherapy Out-patient Clinics | ICGs assisting with ADL<br>Mean age: 39.2 years<br>Age range: 17 to 74 years<br>Sex: Female 76 (48.4%)<br>Employment: Employed: 105 (66.9%);<br>Unemployed: 52 (33.1%) | 157 | Haemorrhagic stroke 61 (38.9%)<br>Ischaemic stroke 96 (61.1%) | Children: 75 (47.8%)<br>Spouse: 45 (28.7%)<br>Children: 21 (13.4%)<br>other significant others: 16 (10.2%) | At least 1 month | Below 6 hours: 57 (36.3%)<br>6–12 hours: 82 (60.5%)<br>> 12 hrs: 18 (11.4%) |
| Gawulayo (2021) (46) | South Africa | Not reported | Family Caregiver assisting with ADL<br>Mean age: 48 years<br>Age range: 33 to 84 years<br>Sex: Female 5 (62.5%)<br>Employment: Employed: 2 (25%); Unemployed: 6 (75%) | 8 | Stroke | Tertiary: 1 (12.5%)<br>Secondary: 7 (87.5%) | At least 6 months | Not reported |
| Esmaili (2018) (63) | Tanzania | Not reported | Caregiver children with cancer<br>Mean age: Not reported<br>Age range: Not reported<br>Sex: Female 11 (55%)<br>Employment: Not reported | 20 | Hepatoblastoma (3), Kaposi's sarcoma (3), Osteosarcoma (3), Wilms tumor (3), Chronic myelogenous leukemia (2), Nephroblastoma (2), Chondroid sarcoma (1), Neuroblastoma (1), Non-Hodgkin lymphoma (1), Retinoblastoma (1) | Not reported | Not reported | Not reported |
| Duru (2021) (36) | Nigeria | Outpatients | Parents of children with Congenital Heart Disease (CHD)<br>Mean age: Not reported<br>Age range: Not reported<br>Sex: Not reported<br>Employment: Not reported | 121 | Acyanotic congenital heart diseases (79.3%), Cyanotic congenital heart (20.7%) | Mothers 84.3%;<br>Guardians 10.7%;<br>Fathers 5.0% | At least 3 months | Not reported |
| Dhada (2019) (47) | South Africa | Outpatients | ICGs of children<br>Mean age: 38 years<br>Age range: 20 to 67 years<br>Sex: Female 14 (100%) | 14 | Diabetes Mellitus | Mother: 9 (64%)<br>Aunt: 3 (22%)<br>Grandmother: 2 (14%) | • The mean duration of each caregiver's experience of caring for a child | Not reported |
|  |  |  | Employment: Employed: 2 (14%); Unemployed: 12 (86%) |  |  |  | with DM was 3.4 (±2.6) years, with a range between 5 months and 10 years<br>• Diabetes caregiving years (total): 47.4 |  |
| Dawson (2020) (67) | Ghana | Inpatient | Primary family caregivers<br>Mean age: 40 years<br>Age range: <30 to >50 years<br>Sex: Female 120 (76.9%)<br>Employment: Employed: 130 (83.3%)<br>Unemployed: 26 (16.7%) | 156 | Lymphoma | Auntie/Uncle: 13 (8.3%)<br>Parent: 108 (69.2%)<br>Grandparent: 19 (12.2%)<br>Sibling: 16 (10.3%) | at least 1 month | Mean care hours/ month 52.3<br>About 56.8 and 54.0 hours per month are spent by carers in the private sector and self-employed (respectively) assisting patients with daily activities. Average time spent by those unemployed is 53.3 hours. |
| Bessa (2012) (48) | Togo | Not reported | ICGs<br>Mean age: 43 years<br>Age range: 30 to 65 years<br>Sex: Female: 14 (82%)<br>Employment: Employed: 7 (41%) Unemployed: 10 (59%) | 17 | Cancer (Not specified) | Siblings: 5 (29%)<br>Spouse: 4 (24%)<br>Children: 3 (18%)<br>Parent: 2 (12%)<br>Relatives: 2 (12%)<br>Friends: 1 (6%) | 9 (53%) had been caring for a loved one between 4 and 12 months prior to the study |  |
| BeLue (2018) (59) | Senegal | Outpatients | Family caregiver<br>Mean age: 39.7 years<br>Age range: 20 to 92 years<br>Sex: Female: 10 (52.6%)<br>Employment: Not reported | 19 | Diabetes mellitus | Children: 7 (36.8%)<br>Spouse: 4 (21.1%)<br>Sibling: 2 (10.5%)<br>Others: 6 (31.6%) | Not reported |  |
| Bekui (2023) (64) | Ghana | Not reported | Family caregiver of a child below 12 years<br>Mean age: 39.7 years<br>Age range: 30 to 69 years<br>Sex: Female 3 (20%)<br>Employment: Employed: 12 (80%) Unemployed: 3 (20%) | 15 | Childhood cancers (Not specified) | Not reported | at least 3 months |  |
| Akpan-Idiok (2014) (37) | Nigeria | Not reported | ICGs<br>Mean age: 35.9 years<br>Age range: <30 to 70 years<br>Sex: Female 132 (62.9%)<br>Employment: Employed 69 (32.9%)<br>Unemployed: 141 (67.1%) | 210 | Cancer (Not specified) | Children: 132 (62.9%)<br>Spouse/partner: 43 (20.5%)<br>Sibling: 21 (10%)<br>Friend: 14 (6.7%) | Not reported |  |
| Adejoh (2021) (9) | Nigeria, Uganda, & Zimbabwe |  | ICGs<br>Mean age: 37 years<br>Age range: 19 to 75 years<br>Sex: Female 24 (50%)<br>Employment: Not reported | 48 | Cancer (Not specified) | Sibling 18 (37.50%)<br>Son/daughter 15 (31.25%)<br>Spouse: 11 (22.92%)<br>Parent 5 (8.33%) | Not reported |  |
| Abba (2022) (38) | Nigeria | Outpatients | ICGs<br>Mean age: 28.1 years | 218 | Stroke | Not reported | 1 – 26 weeks: 204 (93.6%)<br>27 – 52 weeks: | Full-time: 139 (63.8%)<br>Part-time: 79 (36.2%) |
|  |  |  | Age range: 18 to 57 years<br>Sex: Female 132 (60.6%)<br>Employment: Unemployed 83 (38.1%)<br>Employed 135 (61.9%) |  |  |  | 12 (5.5%)<br>53 - 78 weeks: 1 (0.5%)<br>79 - 104 weeks: 1 (0.5%) |  |
| Yousif (2022) (51) | Sudan |  | Parents of children with diabetes<br>Mean age: Not reported<br>Sex: Not reported<br>Employment: Not reported | 150 | Diabetes Mellitus | Fathers and mothers (Proportion not given) | Not reported |  |
| Ogunyemi (2021) (39) | Nigeria | Not reported | Primary ICGs<br>Mean age: Not reported<br>Age range: ≤18 to 60 years<br>Sex: Female 223 (57.9%)<br>Employment: Employed: 231 (60%)<br>Unemployed: 154 (40%) | 385 | Cancer (Not specified) | Parent: 133 (34.6%)<br>Sibling: 93 (24.2%)<br>Spouse: 61 (15.8%)<br>Child: 32 (8.3%)<br>Others: 66 (17.1%) | ≤ 6 months: 125 (43.4%)<br>7 - 12 months: 123 (42.7%)<br>≥ 13 months: 40 (13.9%)<br>Median = 12 (±12.6) | 1 - 5 hours: 204 (53.0%)<br>6 - 11 hours: 132 (34.3%)<br>≥ 12 hours: 49 (12.7%)<br>Median = 6 (±5.8) |
| Malangwa (2022) (58) | Tanzania | Both in and outpatients | Caregivers with children confirmed with cancer.<br>Mean age: Not reported<br>Age range: 18-60 years<br>Sex: Female 70 (72.9%)<br>Employment: Employed: 44 (45.8%)<br>Unemployed: 52 (54.2%) | 96 | Cancer (Not specified) | Father/Mother: 79 (82.3%)<br>Brother/Sister: 8 (8.3%)<br>Aunt/Uncle: 3 (3.1%)<br>Grandmother/father: 6 (6.3%) |  |  |
| Ababacar (2022) (55) | Senegal |  | ICGs<br>Mean age: 42.6<br>Age range: 22 to 75 years<br>Sex: Female 32 (54.2%)<br>Employment: Employed: 42 (70%) Unemployed: 18 (30%) | 60 | Stroke | Children (50%)<br>Spouses (25%)<br>Siblings (13.33%)<br>Nephews/nieces (11.66%) |  |  |

### Quality of the included studies

Overall, the included articles were of good quality as shown in Table 2. More than three-quarters (83%) of these articles met at least four of the five criteria. In two articles (33,55), only two of the five criteria were met. The weaknesses of the quantitative articles were predominantly related to the failure to report or justify the sampling strategy, sample size and risk of nonresponse bias. All the qualitative articles met at least four of the Mixed Methods Appraisal Tool (MMAT) criteria except one (25) where three of the five criteria were met.

**Table 2.** Quality Appraisal of the reviewed articles.

| Author, year | S1 | S2 | Quality scores |  |  |  |  |
| --- | --- | --- | --- | --- | --- | --- | --- |
|  |  |  | 1.1 | 1.2 | 1.3 | 1.4 | 1.5 |
| **QUALITATIVE STUDIES** |  |  |  |  |  |  |  |
| Walubita (2018) (12) | Yes | Yes | 1 | 1 | 1 | 1 | 1 |
| Thomas (2008) (40) | Yes | Yes | 1 | 1 | 1 | 0 | 1 |
| Tchokote (2020) (25) | Yes | Yes | 1 | 0 | 1 | 1 | 0 |
| Owoo (2022) (54) | Yes | Yes | 1 | 1 | 1 | 1 | 1 |
| Owoo (2022) (53) | Yes | Yes | 1 | 1 | 0 | 1 | 1 |
| Mthembu (2016) (41) | Yes | Yes | 1 | 1 | 1 | 0 | 1 |
| Mlaba (2021) (43) | Yes | Yes | 1 | 1 | 1 | 0 | 1 |
| Mlaba (2020) (42) | Yes | Yes | 1 | 1 | 1 | 1 | 0 |
| Mensah (2021) (66) | Yes | Yes | 1 | 1 | 1 | 1 | 1 |
| Masuku, (2018) (44) | Yes | Yes | 1 | 1 | 1 | 1 | 1 |
| Masika (2020) (62) | Yes | Yes | 1 | 1 | 1 | 1 | 1 |
| Maree (2018) (11) | Yes | Yes | 1 | 1 | 1 | 1 | 1 |
| Khondowe (2007) (68) | Yes | Yes | 1 | 1 | 1 | 1 | 1 |
| Ketlogetswe (2022) (45) | Yes | Yes | 1 | 1 | 1 | 1 | 0 |
| Gertrude (2019) (21) | Yes | Yes | 1 | 1 | 1 | 1 | 1 |
| Gawulayo (2021) (46) | Yes | Yes | 1 | 1 | 1 | 1 | 1 |
| Esmaili (2018) (63) | Yes | Yes | 1 | 1 | 1 | 1 | 0 |
| Dhada (2019) (47) | Yes | Yes | 1 | 1 | 1 | 1 | 1 |
| Bessa (2012) (48) | Yes | Yes | 1 | 1 | 1 | 1 | 1 |
| BeLue (2018) (59) | Yes | Yes | 1 | 1 | 1 | 0 | 1 |
| Bekui (2023) (64) | Yes | Yes | 1 | 1 | 1 | 1 | 1 |
| Adejoh (2021) (9) | Yes | Yes | 1 | 1 | 1 | 1 | 1 |
| **NRT QUANTITATIVE STUDIES** | **S1** | **S2** | **3.1** | **3.2** | **3.3** | **3.4** | **3.5** |
| Serfontein (2019) (26) | Yes | Yes | 1 | 1 | 0 | 1 | 1 |
| Scheffler (2019) (27) | Yes | Yes | 1 | 0 | 1 | 1 | 1 |
| **QUANTITATIVE DESCRIPTIVE STUDIES** | **S1** | **S2** | **4.1** | **4.2** | **4.3** | **4.4** | **4.5** |
| Yusuf (2011) (29) | Yes | Yes | 0 | 0 | 1 | 1 | 1 |
| Wassie (2021) (56) | Yes | Yes | 1 | 1 | 1 | 1 | 1 |
| Vincent-Onabajo (2018) (30) | Yes | Yes | 1 | 1 | 1 | 1 | 1 |
| Onyeneho (2021) (31) | Yes | Yes | 1 | 1 | 1 | 1 | 1 |
| Okeke (2020) (32) | Yes | Yes | 1 | 1 | 1 | 1 | 1 |
| Ohaeri (1999) (33) | Yes | Yes | 0 | 0 | 1 | 0 | 1 |
| Ogunmodede (2019) (34) | Yes | Yes | 1 | 1 | 1 | 1 | 1 |
| Muriuki (2023) (50) | Yes | Yes | 1 | 1 | 1 | 1 | 1 |
| Muliira (2019) (57) | Yes | Yes | 0 | 1 | 1 | 0 | 1 |
| Marima (2019) (49) | Yes | Yes | 1 | 1 | 1 | 1 | 1 |
| Kitoko (2022) (52) | Yes | Yes | 1 | 0 | 1 | 0 | 1 |
| Katende (2017) (60) | Yes | Yes | 1 | 0 | 1 | 1 | 1 |
| Jones (2012) (28) | Yes | Yes | 1 | 0 | 1 | 1 | 1 |
| Gbiri (2015) (35) | Yes | Yes | 0 | 0 | 1 | 1 | 1 |
| Duru (2021) (36) | Yes | Yes | 1 | 1 | 1 | 1 | 1 |
| Dawson (2020) (67) | Yes | Yes | 1 | 1 | 1 | 1 | 1 |
| Akpan-Idiok (2014) (37) | Yes | Yes | 1 | 0 | 1 | 0 | 1 |
| Abba (2022) (38) | Yes | Yes | 0 | 1 | 1 | 1 | 1 |
| Yousif (2022) (51) | Yes | Yes | 1 | 0 | 1 | 1 | 1 |
| Ogunyemi (2021) (39) | Yes | Yes | 1 | 1 | 1 | 1 | 1 |
| Malangwa (2022) (58) | Yes | Yes | 1 | 1 | 1 | 1 | 1 |
| Ababacar (2022) (55) | Yes | Yes | 0 | 0 | 1 | 0 | 1 |
| **MIXED METHODS STUDIES** | **S1** | **S2** | **5.1** | **5.2** | **5.3** | **5.4** | **5.5** |
| Mekonnen (2020) (61) | Yes | Yes | 1 | 1 | 1 | 0 | 1 |

*MMAT* Mixed Methods Appraisal Tool [*NRT* Non-randomised Trial, *S1*—Are there clear research questions? *S2*—Do the collected data allow to address the research questions? 1.1—Is the qualitative approach appropriate to answer the research question? 1.2—Are the qualitative data collection methods adequate to address the research question? 1.3—Are the findings adequately derived from the data? 1.4—Is the interpretation of results sufficiently substantiated by data? 1.5—Is there coherence between qualitative data sources, collection, analysis and interpretation? 3.1—Are the participants representative of the target population? 3.2—Are measurements appropriate regarding both the outcome and exposure/intervention? 3.3—Are there complete outcome data? 3.4—Are the confounders accounted for in the design and analysis? 3.5—During the study period, is the intervention/exposure administered as intended? 4.1—Is the sampling strategy relevant to address the research question? 4.2—Is the sample representative of the target population? 4.3—Are the measurements appropriate? 4.4—Is the risk of 0nresponse bias low? 4.5—Is the statistical analysis appropriate to answer the research question? 5.1—Is there an adequate rationale for using a mixed methods design to address the research question? 5.2—Are the different components of the study effectively integrated to answer the research question? 5.3—Are the results adequately brought together into overall interpretations? 5.4—Are divergences and inconsistencies between quantitative and qualitative results adequately addressed? 5.5—Do the different components of the study adhere to the quality criteria of each tradition of the methods involved?], *Y* Criteria satisfied, *N* Criteria not satisfied. 1—Criteria satisfied, *0*—Criteria not satisfied

### Participants characteristics

In this review, two (4.3%) of the 47 articles had samples with paid caregivers forming part of the population, although in small proportions. That is, they made up 8.8% (7/80) of the sample in one article (27) and 1% (1/100) in the second article (34). Similarly, caregivers whose care recipients had died formed part of the sample in two articles (42,43). For each study, these caregivers accounted for 15% of the respective samples. There was no study with a comparison group. Also, ten articles (29,30,32,34,38,49,52,56–58) (41.7% of the quantitative articles) noted that they excluded ICGs who had any of the outcomes of interest prior to assuming the caregiving role.

The majority of caregivers were either parents, siblings, children, spouses or other relatives of the patient and their ages ranged from ten (31) to ninety-two years (59). The proportion of non-familial caregivers was less than 10% in each of the seven (14.9%) articles (27,32,34,37,45,48,60) where they were part of the sampled populations. In 19% (9/47) of the articles (12,36,47,51,58,61–64), the participants were caregivers of children with chronic diseases. More than half of the articles (53.2%, 25/47) focused on caregivers of people with cancers (9,11,12,29,31,33,37,39,42,43,45,48,53,54,56–58,60–67). The rest of the articles studied caregivers of people with cardiovascular diseases (21,25–28,30,32,35,36,38,40,44,46,49,52,55,68) (36.2%, 17/47) and diabetes mellitus (34,47,51,59) (8.5%, 4/47). There was no study with caregivers of people with chronic obstructive pulmonary disease. Generally, there were more female participants than male participants. However, in just five (10.6%) of these papers (29,35,56,64,66), the proportion of females did not reach at least half of the total participants. The characteristics of the included studies and participants are summarised in **Table 1**.

### Activities performed by ICGs

The roles performed by caregivers were described in two-fifths (20/47) of the included articles. With the exception of one (45), these articles listed the various things caregivers did. In this article, the authors simply stated that performed activities of daily living were demanding and exhausting but did not specify what tasks they were.

In this review, we have grouped informal caregivers’ activities into three activities.

#### i. Basic activities of daily living

These are personal care tasks that are important for one’s everyday living. The ICGs played a critical role in supporting care recipients with personal care activities like bathing and dressing (9,11,21,25,35,40,41,48,54,66,68), toileting (21,48) and feeding (21,25,64). They also assisted with functional mobility activities that support the patient to move in and out of bed, repositioning, ambulating and moving up/down the stairs (21,25,35,40,48).

#### ii. Instrumental activities of daily living

These activities were quite diverse and included caring for the patient’s children (40), shopping for the patient (11,68) and doing domestic tasks (house chores) like room cleaning, making meals, laundry and washing utensils (9,11,25,36,40,46,48,54,59,66). The caregivers were actively involved in the health management of their sick loved ones by managing medical appointments and transporting the patient to hospital (9,11,21,25,36,42,46,47,53,59,62,64,68), communicating with health workers on the patient’s behalf (9), paying medical expenses (9,21,35,36,42,47,48,59,62,63), providing company and psychological support to patient (9,21,59,62) and monitoring the patient’s symptoms (11,47,66).

#### iii. Medical activities

The ICGs performed activities that would ideally require a certain level of medical training, experience or knowledge. Eight articles (9,11,21,25,40,41,59,66) highlighted specific medical procedures that were done by the ICGs. These included changing colostomy bags, wound dressing, helping patient use oxygen breathing apparatus and administering both oral and injectable medications for the patient. Despite performing these tasks, the ICGs acknowledged their lack of knowledge and skills. This was clearly articulated by one caregiver: “*To be honest I cannot say I have been doing a great job of caring for my father.…it is just a ‘trial and error’ situation. It feels like I am not doing enough. Somehow, I feel like giving up…*’(45).

### Reasons/motivation to provide care

The reasons for taking up caregiving roles were recorded in six (12.8%) papers (11,21,25,45,48,64), all of which were qualitative studies. From their submissions, it was clear that the decision to provide informal care depended on two things; how related or connected the ICG was to the patient and the ICG’s level of responsibility (social obligation) over the patient.

#### i. Relationship between the caregiver and the patient

Regarding the relationship (or connectedness) to the patient, ICGs narrated how they provided care because of the love that existed in their relationship either as family members, neighbours or friends (21,48). Because of this bond (and feeling of belonging), they willingly volunteered and often considered themselves the most suitable persons to care for their loved ones. The closeness of the relationship between the caregiver and recipient is a strong motivator for becoming a caregiver. This view was echoed by a Togolese caregiver who was quoted saying, “*The problem is that nobody can take care of my mother as I am doing it. If something does not belong to you, you treat it differently.*” Another participant in the same study added, “*I see myself in a better position to do the care work because I know her very well.*” (48).

#### ii. Level of responsibility the caregiver has over the patient

Quite relatedly, the ICGs also viewed their decision to care as a form of responsibility or a social obligation (11,25,45,48,64). They naturally took on the role and were as committed in the work as they would in any other professional job. They considered caring as a duty they were obliged to perform against all odds for those they were socially expected to support. One mother explained how she had to take her older child (with cancer) to the hospital despite her own poor health: “*I remember there was a time I sat in a chair by the bedside throughout the night and my legs got swollen…it was not easy, but I passed through all that. When I gave birth on Thursday, the following Tuesday I had to bring my sick son to the hospital for review…I had to do it.*” (64)

### Impact of caregiving on the caregiver

All studies had an aspect of the impact (health or economic) caregiving had on the carers. In this section, results on how the impact was measured, positive impact of caregiving, economic impact of caregiving and health impact of caregiving was provided. These are also summarised in Table 3

**Table 3.**
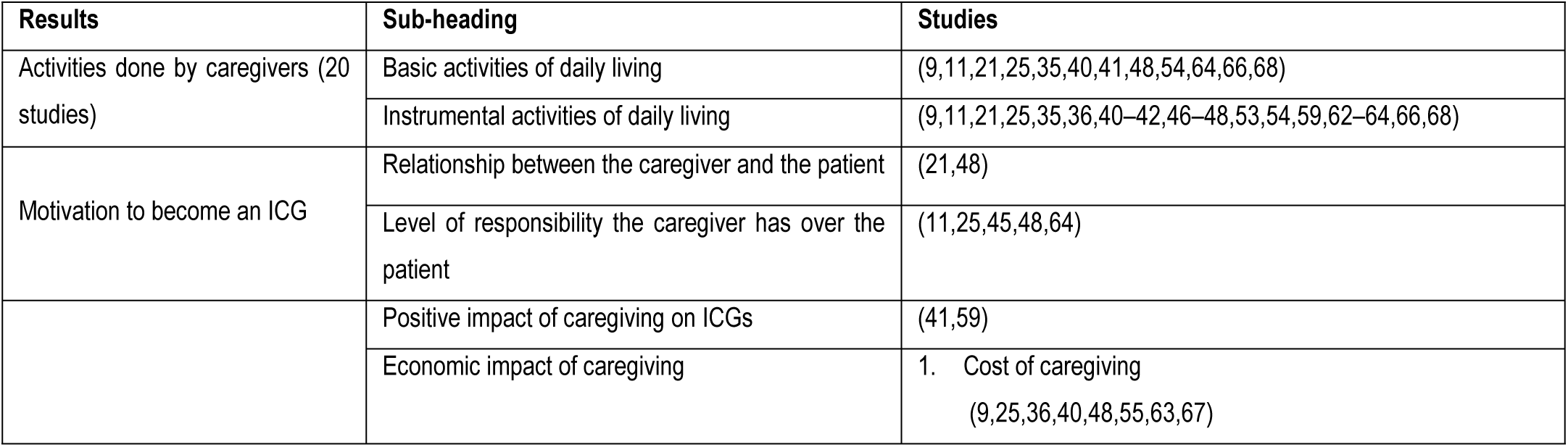
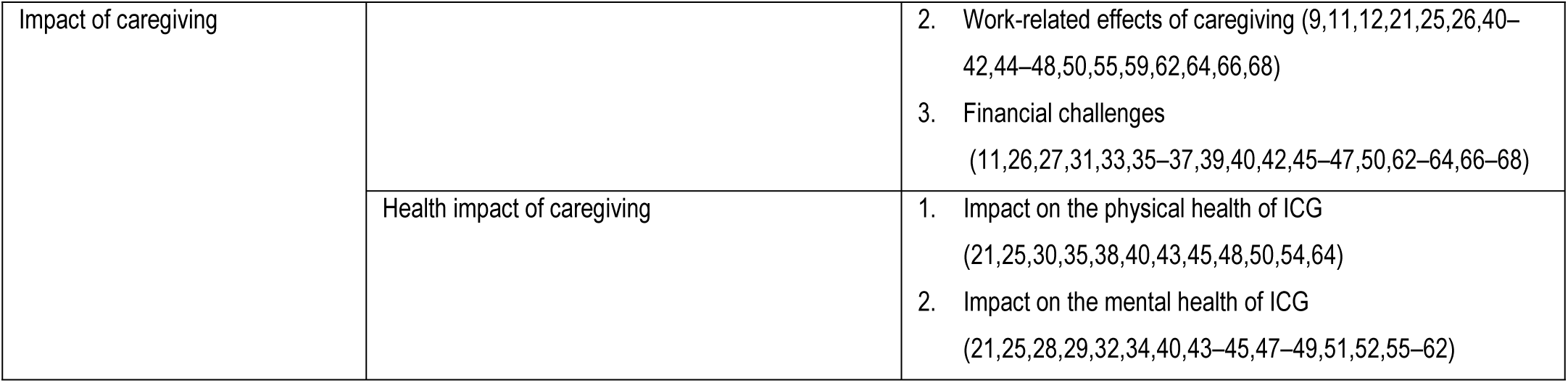
Table showing the results identified in the included articles.

#### Measurement of the impact of caregiving

From the findings, about half of the articles (22/47) used standardised tools/instruments to determine the impact of caregiving. It was only the qualitative papers and one quantitative paper (36) where no standardised instrument was used in measuring the impact of caregiving. The tools (in their original or modified form) used include, the Zarit Burden Interview (ZBI) questionnaire (29,31,32,34,35,37,39,52,55,67), the Hospital Anxiety and Depression Scale (HADS) (28,52,57,60), Caregiver strain index (CSI) (26,27,35,50), Nordic Musculoskeletal Questionnaire (NMQ) (30,38), General health questionnaire (GHQ) (29,33,34), Patient Health Questionnaire-9 (PHQ-9) for Depression (56) Caregiver Reaction Assessment scale (CRA) (57), Beck’s depression inventory (BDI) scale (61), Shona Symptoms Questionnaire (SSQ) (49), The modified Rankin scale (mRS) (52), Depression, Anxiety, Stress Scale-21 Items (DASS-21) (51) EUROHIS QoL (67), Modified version of the Frankfurter Befindlichkeit Skala (FBS) (33) and Hopkins Symptom Checklist (HSCL-25) tool (58). Two studies (36,67) used cost of illness approaches to estimate the direct and indirect costs incurred by the caregivers who supported people with chronic diseases.

#### Positive impact of caregiving

Although the focus of most articles was on the negative impact of caregiving, two articles (41,59) in particular had caregivers who described how rewarding the activity was to them. They commented on how the situation strengthened the bond with the person being cared for, thus enabling them to develop the dedication and emotional strength to continue caring. Furthermore, the practice of caring became a strong motivation for improvement of their own health (59). Whenever they saw how their loved ones suffered, they felt energized to do whatever was necessary to avoid getting the same disease. They became motivated to adopt a healthier lifestyle that included having regular medical check-ups, doing more exercises and taking healthier meals.

Because of their involvement in the day-to-day management of the care recipients, they acquired skills and knowledge to protect themselves from acquiring the disease and to fulfil the care needs better. For example, one caregiver who was always there whenever the doctor came to treat his wife stated that, “*my experience helps a lot to avoid diabetes in my own life. What the doctor says to my wife, I also apply to my own life because that is good for everyone, not just the diabetic patients.*” (59)

#### Economic impact of caregiving

There were three key findings under economic impact, and these were the cost of caregiving, the work-related consequences of caregiving and the subsequent financial challenges arising from caregiving.

##### i. Direct costs of caregiving

Surprisingly, only 4 articles (36,48,55,67) provided information on the cost of caregiving on the caregiver. Of these, only 2 articles gave an estimate of the overall cost of providing informal care. According to the studies by Dawson *et al* (67) and Duru *et al* (36), the monthly mean cost of caregiving for each patient was US$440.32 (SD: US$205.75) and US$ 20.36 ± US$ 27.82, with direct costs accounting for 97% and 81% of the total costs respectively. The only information on the costs of care that *Bessa et al* provided was the average monthly money (US$ 66 monthly (range = US$ 21 – US$ 213)) spent by ICGs on their loved ones with cancer (48).

Two articles (36,55) discussed the monetary value of the productivity loss incurred by ICGs while providing care. The mean productivity loss incurred by caregivers in Duru *et al*’s study (36) was $53.42 ± $88.12 (range US$1.38 to US $ 575.00) while those in the study by Ababacar *et al* (55) experienced a financial loss of 206.92 USD (SD: 583.16 USD) (range: 0 to 3103.44 USD).

##### ii. Social cost of caregiving

Of note, are the 4/47 articles (9,25,40,63) that commented on the social cost associated with informal caregiving. Opportunity cost refers to the cost of foregoing other rewarding activities in order to support a sick loved one. As one ICG put it, caregiving “*restricts our lives*” (40) giving them little room for engagement in other activities. A female ICG in Cameroon commented on the cost involved in letting go of her previously planned activities. She stated, “*Since my mother is in this state, […] I am obliged to suspend my participation in the church every morning as I did before because it is no longer possible! I had to review all my activities because I had to find time for my mother! It’s painful but I do it!*” (25).

While some caregivers had to let go of their regular leisure activities (25), others especially the students ended up missing attending classes at school (40).

##### iii. Work-related consequences of caregiving

In 21 (44.7%) studies, there is a clear demonstration of how caregiving affected all aspects of one’s work life (regular source of income). Since caring often required one’s physical presence, the caregivers not only used up all the available work leave (including taking leave without pay) but they were also frequently absent (without permission) from work. This resulted into being queried, warned, sanctioned and in other cases dismissed (9,11,48,64,66). In the study by Ababacar et al. (2022) (55) the caregivers were off work for an average of 23.86 days (SD 64.53 days) per year with extremes ranging from 0 to 365 days.

The participants admitted that they struggled to maintain a balance between work and caregiving roles (9,11,21,41,48,59,66,68). This made them less efficient (productive) at work and unable to meet several deadlines because they frequently arrived late for work, spent less time doing work and struggled to concentrate while at work (12,25,42,48,66). This situation was well explained by the caregiver who said, “*There was a piece job that I had in Durban, it was affected because I had to be here in Pietermaritzburg to take care of her [my mother]. I couldn’t stay [at work] for the week and come back on the weekend or month end, I had to keep coming back here [to check on my mother].*” (42)

As such, they had to make work-related adjustments that would allow them to attend to their sick loved ones too. Part of the adjustments included getting a flexible work schedule that allowed them to work either at night or from home (26,44,50,55) and changing to temporary self-employment or to jobs that are flexible and closer to the patient’s home (42,46,55). In cases where the caregivers could not maintain both work and caregiving roles, they either voluntarily resigned or were dismissed from work (9,11,12,21,40–42,45–48,55,62,64,66).

##### iv. Financial challenges arising from caregiving

There were 21 (44.7%) articles in which caregivers reported experiencing financial challenges. First, they stated that they lost money and their sources of income through covering high medical expenses and loss of jobs (31,33,36,42,45,63,64). Their response to such challenges included taking loans to get money to care for the patient (33,63,64,66,67), seeking additional financial support from relatives and institutions like churches (62,67), sale of family properties (62,64,66) and requesting for premature discharge from the hospitals (63).

In cases where resources to cover the expenses were inadequate, the ICGs compromised their standard of living, forewent paying house rent, paying their children’s school fees as well as buying food and clothes in order to meet their patient’s care needs (9,63). As a result, some were reportedly thrown out of their house due to non-paid rent.

Overall, caregivers felt that caring was financially burdensome causing them to be in a much less satisfactory financial status than before taking up the role (11,26,27,35,37,39,40,42,46,47,50,66,68). They described their financial situation as being unstable, strenuous and hard, and they often became anxious whenever they thought about where to get more money to continue with their role (35,46,47). Quantitatively, the prevalence of caregivers experiencing financial strain ranged from 37.9% to 100% (26,27,37,50).

#### Health Impact of caregiving

Caregiving affected the physical health and mental health of the carers as stated below.

##### i. Physical health consequences of caregiving

This was found in twelve articles (21,25,30,35,38,40,43,45,48,50,54,64). The development of new medical conditions and or worsening of existing health conditions among the ICGs affected their physical health and reduced their ability to adequately care (35,64). Some attributed this to lowering of the body’s immunity (40) that occurred while providing care. They stated that they developed new medical conditions such as hypertension, headache, body pains and swelling of the feet during the period they provided care to the patient (21,45,64). One caregiver even narrated a life-threatening incident that necessitated hospitalization “*…after collapsing while fetching water for the sick son*” (64). There were also reports of the worsening of the caregivers’ own health condition (21,30,38,45,48,54,64). The articles show how those with musculoskeletal complications such as pain in upper and lower back, pain in different joints (shoulder, wrist, hips, buttocks, knees, ankle and elbow joint), and pain in the thighs, neck and feet complained of deteriorating health. This was well articulated by a caregiver whose waist problem worsened while caring for her husband. She stated; “*Caring for my husband is not easy; I have a waist problem, and I find myself always very tired. The waist problem was there but was not as serious as it has become now, all because of the role I currently play*” (54).

The caregivers in 10 studies experienced significant lifestyle changes during their work. They felt that their sleep was disturbed because of actual/anticipated care demands, and severity of the condition of the person they cared for (64). They experienced disruption from sleep, feeling of lack of enough sleep, feeling like they are deprived of sleep as well as short episodes of sleep (25,43,45,50,54,64). Another way caregiving affected their lifestyle was through change in eating habits. This involved either changing the meal time in order to incorporate the caregiving demands/activities, eating the patient’s left over foods or missing the meals due to the lack of money/ time (54,64). A male carer commented on his changed lifestyle by saying, “*It [life] has changed because I hardly sleep now, I just think that I can’t sleep I have to constantly check on him*.” (43)

##### ii. Mental health consequences of caregiving

The act of caregiving had a profound effect on the mental wellbeing of care providers as stated in more than half (23) of the articles (21,25,28,29,32,34,40,43–45,47–49,51,52,55–62).

Although most of these articles described the different types of mental health challenges, five (29,34,49,55,58) did not specify as they only used the terms “psychological” and “psychiatric” challenges. Using GHQ (29,34), SSQ (49), HSCL-25 (58) and ZBI (29,34,55) tools, the reported prevalence of psychological/psychiatric challenges among caregivers ranged from 35% to 66.7%.

The caregivers experienced mild to severe forms of depression as was assessed using HADS (28,52,57,60), ZBI (32), PHQ-9 (56), BDI (61) and DASS-21 (51) tools. From these eight quantitative studies, the prevalence of depression ranged from 26% to 72.4%. In addition, there were two qualitative studies with ICGs who reported feeling depressed because of the caregiving work (25,40). Some got to an extent of being treated for depression as explained by this South African ICG: “*… I also get sick often. I have continuous lung infections from stress. I’m on antidepressants as well*.”

Fear, stress and anxiety is another mental health related challenge that caregivers in 9 studies (28,45,47,51,52,57,59,60,62) reportedly experienced. There was no quantitative paper in which fear or stress was investigated. Instead, five quantitative papers employing HADS (28,52,57,60) and DASS-21 (51) tools found that the prevalence of anxiety among ICGs ranged from 21.6% to 45%. Sometimes, the ICGs thought about the likelihood of acquiring the same illness their loved ones have or death of those they care-for. Such thoughts caused fear, stress and anxiety among the ICGs (45,59). This challenge was most felt in the initial stages of caring for the patient, when there was a lot of work to be done and when the activities were complex (47). While one caregiver hinted at how hard some caregiving activities can sometimes be by saying, *“the hardship is just injecting the child part, it’s not something that’s easy—it’s not easy – because sometimes you find that the child does not want to be injected…when her sugars are high and you try to inject her – she says ‘you’re injecting me a lot’.”*(47)

Another expressed the fear that comes with such activities: *“… The family members you stay with do want to assist with injecting the child but they are scared – so sometimes you’re the only one who’s taking care of the child.”* (47)

Five quantitative articles (28,51,52,57,60) explored the prevalence of anxiety among caregivers and found it to be in the range of 21.6% to 45%.

Caregiving was also described as an emotionally challenging activity by participants in four studies (25,44,45,48). They used phrases such as being morally sick, emotionally painful, heartbreaking, emotionally touched, burdens me, disturbed and hopelessness to show their reaction to caring for the sick or the diagnosis given. The extent of suffering that caregivers observed their loved ones go through was enough to create feelings of preferring to “*isolate [oneself] to cry”* and develop thoughts of “*not … living anymore*” (44,48). Caregivers confessed that such reactions also followed the sudden announcement (by the medical team) of the type disease their loved one was suffering from (25). One participant whose husband had been diagnosed with cancer recalled such a moment: “*It was painful, wena [‘you’ – exclamation]. You … you know that person is a breadwinner and everyone in his family is looking up to him. He is supporting everyone. Then it was painful, but I think that I am recovering now.*” (44)

## Discussion

In this review, we found a total of 47 qualitative and quantitative studies from across the sub-Saharan African region that were published over a period of more than two decades.

### Reasons of becoming an ICG

These studies show that a relationship that is strengthened by love and the sense/feeling of responsibility over an individual appear to be the most important factors in the decision to provide care. This finding is consistent with those in other reviews with studies from across the world (17,69). In our review, children as young as ten years of age also participated in providing care to people with chronic diseases. This can be explained by how responsibilities/activities are shared or delegated in many African multi-generational homes (29,70). Often, children are assigned the task of providing care for unwell or elderly relatives while adults participate in income generating activities (29,70). Once assigned the tasks, the children take them as their responsibility. The finding from this and other reviews (17,69) may mean that the reasons for becoming caregivers may not be very different across the world even with varying cultures and traditions.

### Activities performed by ICGs

The caregivers, majority of whom were female and were related to the patient, performed a wide range of activities during their caregiving role. Perhaps the most striking finding is that caregivers did not end at performing the common/regular tasks like support with feeding and with personal care needs, but they did complex activities including changing colostomy bags and administering injectable medication which normally require medical skills/training to do well. This rather interesting finding points to shortage of health workers in most settings in SSA. In most cases, the ICGs end up performing these tasks when health professional cannot be accessed (71,72). The implication of such findings is the possibility of ICGs making numerous mistakes while conducting procedures they were not trained to perform and unknowingly endangering themselves and/or those under their care. A recent review conducted in one of the SSA countries had findings similar to ours (16). In their study, the caregivers were mainly females, relatives of the patient and performed most activities as described in our work. The only noticeable difference is that there was no mention of the activities, that we have described as complex. This is possibly because their study either considered diseases (such as HIV) that are rarely complicated by events like surgery which may require changing colostomy bags and wound dressing or the diseases had not yet gotten to a level that required interventions like use of oxygen therapy or administration of injectable medications.

### Impact of caregiving on the ICGs

To measure the impact of caregiving, almost all quantitative studies used a standardised instrument/tool. As shown in the results, a total of 14 different tools were used in the 22 studies to assess the burden of depression, anxiety, stress and financial strain. Even in the studies where the same tool was used, the results were presented differently. For example, in the studies that used Zarit burden Interview (ZBI), two (29,32) reported the overall mean ZBI scores while the rest provided mean scores of each category of burden. This lack of uniformity in the use of measures and presentation of results make it difficult to compare different studies’ results. In this review, ZBI tool was the most used instrument for measuring the impact of caregiving. It was used in 10 out of the 22 articles assessing the health or economic impact using an instrument. This accords with earlier observations from another review which found that ZBI was the most frequently used tool in measuring the burden of caregivers (73).

As discussed in the next three paragraphs, caregiving had both positive and negative effects on the ICGs. As expected, there were more articles focussed on the negative than positive impact of caregiving.

The rewards experienced by informal caregivers in this review can be explained by the post-traumatic growth (PTG) theory which suggests that people facing traumatic or challenging events may experience some positive outcomes from their experience (74). Caring for people, some of whom are at an advanced stage of a chronic disease can be stressful and traumatic due to the highly demanding nature of the caregiving tasks. Such situations often prompt ICGs to do self-reflection leading to the discovery of new and positive insights as well as renewed purpose for living under the current conditions. A number of recent studies have used this theory to demonstrate how caregivers’ challenging experiences resulted in positive outcomes like renewed personal strength to continue in their caring role, strengthening of the relationship with care recipients/family members and taking better care of their own health because they better appreciate life (75–77).

As with other reviews (15,16) conducted in SSA, caregivers with a regular source of income faced work-related challenges such as absenteeism, dismissal from work and low productivity as already listed in the results section. Caregivers’ involvement in extensive physical work like assisting patient with mobility-related activities may result into injuries and exhaustion. Depending on the patient’s care needs and severity of the disease, the caregivers may spend a lot of time with the patient. Under such circumstances, the caregivers will either fail to go to work or arrive late at work leading to reduced productivity and conflicts at work. The exhaustive nature and emotional burden that comes with caring for loved ones also reduces one’s level of concentration and performance while at work. In the end, caregivers are often dismissed or forced to resign from their work. At times, they sought employment in places with work that suited their caregiving responsibilities such as self-employment, temporary employment or jobs with flexible working hours (78). It is also worth noting that a disrupted career/job has the potential of causing significant financial strain on the career, hinder career progression and affect retirement planning. These are likely to persist even after they are no longer providing care.

As would be expected, informal caregivers experienced both physical health and mental health challenges during their work. Several reviews have had similar findings published (14–17). This is consistent with allostatic load theory, in which McEwen and Stellar observed that although stressors usually resulted in a normal physiological response as a body’s protective mechanism, exposure to high and sustained stress such as that experienced by informal caregivers caused an abnormal response that resulted in cumulative wear and tear of the body (79). The allostatic load experienced by caregivers arises from exhaustive physical activities they do, emotional challenges caused by the patient’s condition and logistical stressors like financial strain. Part of the abnormal response includes change in the production and levels of hormones like cortisol and adrenaline which could lead to cardiovascular diseases, weakened immune system, inflammation, metabolic changes (affecting an individual’s appetite) and mental health diseases.

### Limitations of the review

This review had some limitations. First, the second reviewer could only screen a proportion of studies during the title and abstract screening stage and during the full text screening. Also, a single author extracted the data from the included studies with the second reviewer checking the accuracy of the data. A study showed that a complete dual review significantly minimised random errors compared to either single review or limited dual review (80).

Secondly, we were unable to get all the full-texts of the conference abstracts that were included in this review. While the contacts for some authors could not be retrieved, the majority of those contacted either did not respond or did not have the full-texts for their abstracts available. It is therefore possible that some important articles could have been missed.

Furthermore, some studies did not provide adequate information as some data was either unclear or not reported. This includes information on sex, age and well as the impact of caregiving. This limited our ability to extract all the relevant data required to comprehensively analyse the data.

Lastly, most quantitative studies used a cross-sectional study design making it difficult to assess the cause-and-effect relationship, and to understand the changes that occur over time. However, almost half of these articles excluded participants with outcomes of interest prior to starting caregiving.

### Strengths of the review

Part of the strength of this review is in the fact that we used a very broad search strategy to extensively search a good number of relevant databases. We also included non-English articles. These enabled us to capture as many relevant articles and information as possible. The use of a second reviewer improved the reliability and quality of the findings through reducing the errors and biases that could have risen if only one reviewer was involved in the review process.

## Conclusions

The ICGs occupy an essential place in the health care system of most SSA countries and they perform a broad range of tasks, some of which they are ill-prepared or untrained to do. The growing demand for ICGs created by the increasing chronic diseases in the African continent means that health care systems would function better if they are adjusted to prepare ICGs for the caregiving roles, and to support the physical and mental wellbeing of ICGs. Future research should investigate the most suitable tools/instruments for assessing the impact of care on the caregiver.

## Data Availability

All relevant data are within the manuscript and its Supporting Information files

## Supporting Information

**S1 Fig. PRISMA flow diagram of the screening and selection process in this review.**

**S1 File. Search strategy used in the selected databases.**

